# Racial Disparities in Knowledge of Cardiovascular Disease by a Chat-Based Artificial Intelligence Model

**DOI:** 10.1101/2023.09.20.23295874

**Authors:** Oseiwe B. Eromosele, Temitope Sobodu, Olakeyede Olayinka, David Ouyang

## Abstract

**Background:** Patients and their families often explore the information available online for information about health status. A dialogue-based artificial intelligence (AI) language model (ChatGPT) has been developed for complex question and answer. We sought to assess whether AI model had knowledge of cardiovascular disease (CVD) racial disparities, including disparities associated with CVD risk factors and associated diseases.

**Methods:** To assess ChatGPT’s responses to topic in cardiovascular disease disparities, we created six questions of twenty sets, with each set consisting of three questions with changes to text prompt based on differences in patient demographics. Each question was asked three times to assess variability in responses.

**Results:** A total of 180 responses were tabulated from ChatGPT’s responses to 60 questions asked in triplicate to assess. Despite some variation in wording, all responses to the same prompt were consistent across different sessions. ChatGPT’s responses to 63.4% of the questions (38 out of 60 questions) were appropriate, 33.3% (20 out of 60 questions) were inappropriate and 3.3% unreliable (2 out of 60 questions). Of the 180 prompt entries into ChatGPT, 141 (78.3%) were correct, 28 (15.5%) were hedging or indeterminate responses that could not be binarized into a correct or incorrect response, and 11 (6.1%) were incorrect. There were consistent themes in incorrect or hedging responses by ChatGPT, with 91% and 79% of incorrect and hedging responses related to a cardiovascular disease disparity that affects a minority or underserved racial group.

**Conclusion:** Our study showed that an online chat-based AI model has a broad knowledge of CVD racial disparities, however persistent gaps in knowledge about minority groups. Given that these models might be used by the general public, caution should be advised in taking responses at face value.

## Introduction

In obtaining information about cardiovascular diseases, individuals, including health professionals, health professional trainees, and students, explore the information available online and from their healthcare providers. A popular dialogue-based artificial intelligence (AI) language model (ChatGPT), that responds to complex queries interactively was released in November 2022 and has gained much coverage lately, with currently about 100 million users. ^1–3^

The National Institute of Health (NIH) describes health disparities are differences in the incidence, prevalence, mortality, and burden of diseases and other adverse health conditions that exist among specific population groups in the United States. ^4–6^ Ethnicity and race on the other hand are socially constructed categories with meaningful impact in the lives of those defined by these constructs, and how these constructs influence people’s perception of an individual, with an appreciation of the complexity that defines the social construct including prevailing social perceptions, historical policies, and practises. ^4^ Despite the overwhelming evidence that consideration of race, ethnicity and other social determinants of health factors are central and extremely important in addressing health equity; as well as the many strides that have been made to improve healthcare delivery in the US, racial disparities continues has continued to persist in healthcare. ^4,7–10^

In spite of the promise of AI systems in addressing health disparities, they are uniquely positioned to exacerbate health inequities, doing this by reproducing and/or magnifying the biases of their human designers or training data sets in ways that risks hence the need to measure how AI systems potentiate these disparities. ^11–13^ This brings up the need to assess this exacerbation of health disparities by AI systems, allowing for the development of targeted measures to limit and curb this negative impact of AI systems, to guarantee reliability in using these systems especially with their growing use. ^14^ We sought to assess an online chat-based AI model’s (ChatGPT) knowledge of cardiovascular disease (CVD) racial disparities, including disparities associated with CVD risk factors and associated diseases, in what we believe would be the first study to do so.

## Methods

### Prompt Construction

We created a total of sixty questions from twenty question sets, with each set consisting of three questions framed as similar questions around a certain topic in cardiovascular disease disparities totaling thirty questions, each posed to ChatGPT to assess its knowledge of cardiovascular disease racial disparities, including its knowledge of similar disparities on CVD risk factors and associated conditions, that is currently available research data on health disparities and clinical care in general and subspecialty cardiology clinical settings. ^15,16^

### Evaluation and Statistical Analysis

Each question was posed to ChatGPT 3 times to assess for variability in responses. ^17–21^ After each question prompt, ChatGPT was closed and reopened to check for inconsistency and variation in the output. The 180 responses from each of the 3 entries of the 60 questions were recorded, following which all responses obtained were be graded individually assessed as ‘yes’ if the response was correct, ‘hedge’ if the response did not explicitly yield a correct response, and ‘no’ if the response was either wrong or misleading. Based on these ‘yes’, ‘hedge’ and ‘no’ responses, the 60 questions were further graded as ‘appropriate’, ‘inappropriate’ or ‘unreliable’, where the question is graded as ‘appropriate’ if three of the responses in the question contained appropriate and consistent information; ‘inappropriate’ if any of the three responses contained inappropriate or inconsistent information, and ‘unreliable’ if the three responses were inconsistent. ^17^ The responses were assessed and graded by a team of experienced cardiologists and other clinicians with research and/or clinical focus on cardiovascular health disparities. This study was performed in June 2023. This study was not human subjects research and waived from institutional review board review. Statistical analysis was performed in August 2023.

## Results

A total of 180 responses were tabulated from ChatGPT’s answers to 60 questions in triplicate to assess for consistency. All responses to the same prompt were consistent across different question and answer sessions. ChatGPT’s responses to 63.4% of the questions (38 out of 60 questions) were appropriate, 33.3% (20 out of 60 questions) were inappropriate and 3.3% unreliable (2 out of 60 questions). Of the 180 prompt entries into ChatGPT, 141 (78.3%) were correct, 28 (15.5%) were hedge responses and could not be binarized into a correct or incorrect response, and 11 (6.1%) were incorrect. There were consistent themes in incorrect or hedging responses by ChatGPT, with 91% and 79% of no and hedge responses respectively, related to a cardiovascular disease disparity, risk factor or related condition that affects a minority, or underserved racial group. ^5^

Of the 91% of the no responses were related to a minority or underserved racial group, with 45.5% of the responses related to non-White Hispanic Americans, 18.2% related to Asian Americans, and 9.1% related to Cuban Americans, Pacific Islander Americans, and Puerto Rican Americans respectively. On the other hand, 79% of the hedge responses were related to a minority or underserved racial group, with about 32% of the responses related to non-White Hispanic Americans, while 18.2% of the responses were related to African Americans and Asian Americans, respectively. Other hedge responses were related to Puerto Rican Americans at 13.6%, while 9.1% were related to Pacific Islander and Cuban Americans.

CVD racial disparities that yielded hedge responses included heart disease, myocardial infarction, heart failure, acute coronary syndrome, dyslipidemias, metabolic syndrome, body mass index and peripartum cardiomyopathy. On the other hand, ChatGPT yielded incorrect responses to CVD racial disparities on the following topic areas heart disease, stroke, acute coronary syndrome, systemic hypertension, diabetes mellitus, peripheral vascular disease, and metabolic syndrome yielded no responses.

## Discussion

Our study found that a popular online chat-based AI model’s (ChatGPT) knowledge of CVD racial disparities, risk factors and related conditions was appropriate albeit suboptimal as evaluated by cardiologists with clinical and research expertise in cardiovascular health disparities. This exploratory study also highlights some important findings such as the proportion of hedge and no responses affecting groups already categorized as minority or marginalized. ^5^

Also important to highlight are the proportion of important cardiovascular disease conditions that the online AI model provides incorrect responses including acute coronary syndrome, heart failure, peripartum cardiomyopathy, systemic hypertension and metabolic syndrome to mention a few, that already disproportionately affect these minority and vulnerable racial groups. ^22^

Similar to findings by Sarraju et. al. on the appropriateness of CVD prevention recommendations obtained from an AI model, our study similarly showed satisfactory appropriate knowledge as can be seen in, albeit lower at about 64% as shown in Table 1 and Figures 2, 3 and 5, compared to 84% in the study by Sarraju et.al. ^3^

**Table 1.** Graded Responses and Associated Minority Groups. This table above presents the distribution of graded responses categorized as ‘Yes,’ ‘Hedge,’ or ‘No,’ alongside the respective minority groups associated with each response outcome. The table provides an overview of how these responses are distributed among different minority groups, offering insights into variations in response patterns across the study population.

| Results Table |  |  |
| --- | --- | --- |
| Appropriateness Report | Appropriate | 38/60 |
|  | Inappropriate | 20/60 |
|  | Unreliable | 2/60 |
| Individual Responses | Yes | 141/180 (78.3%) |
|  | Hedge | 28/180 (15.6%) |
|  | No | 11/180 (6.1%) |
|  | Hedge responses involving a minority racial group (%) | 22/28 (79%) |
|  | No responses involving a minority racial group (%) | 10/11 (91%) |
| Breakdown of Hedge Responses | Non-White Hispanic Americans | 7/22 (31.8%) |
|  | Asian Americans | 4/22 (18.2%) |
|  | African Americans | 4/22 (18.2%) |
|  | Puerto Rican American | 3/22 (13.6%) |
|  | Pacific Islander American | 2/22 (9.1%) |
|  | Cuban American | 2/22 (9.1%) |
| Breakdown of Incorrect (No) Responses | Non-White Hispanic Americans | 5/10 (50%) |
|  | Asian Americans | 2/10 (20%) |
|  | Cuban American | 1/10 (10%) |
|  | Pacific Islander American | 1/10 (10%) |
|  | Puerto Rican American | 1/10 (10%) |

**Figure 1.**
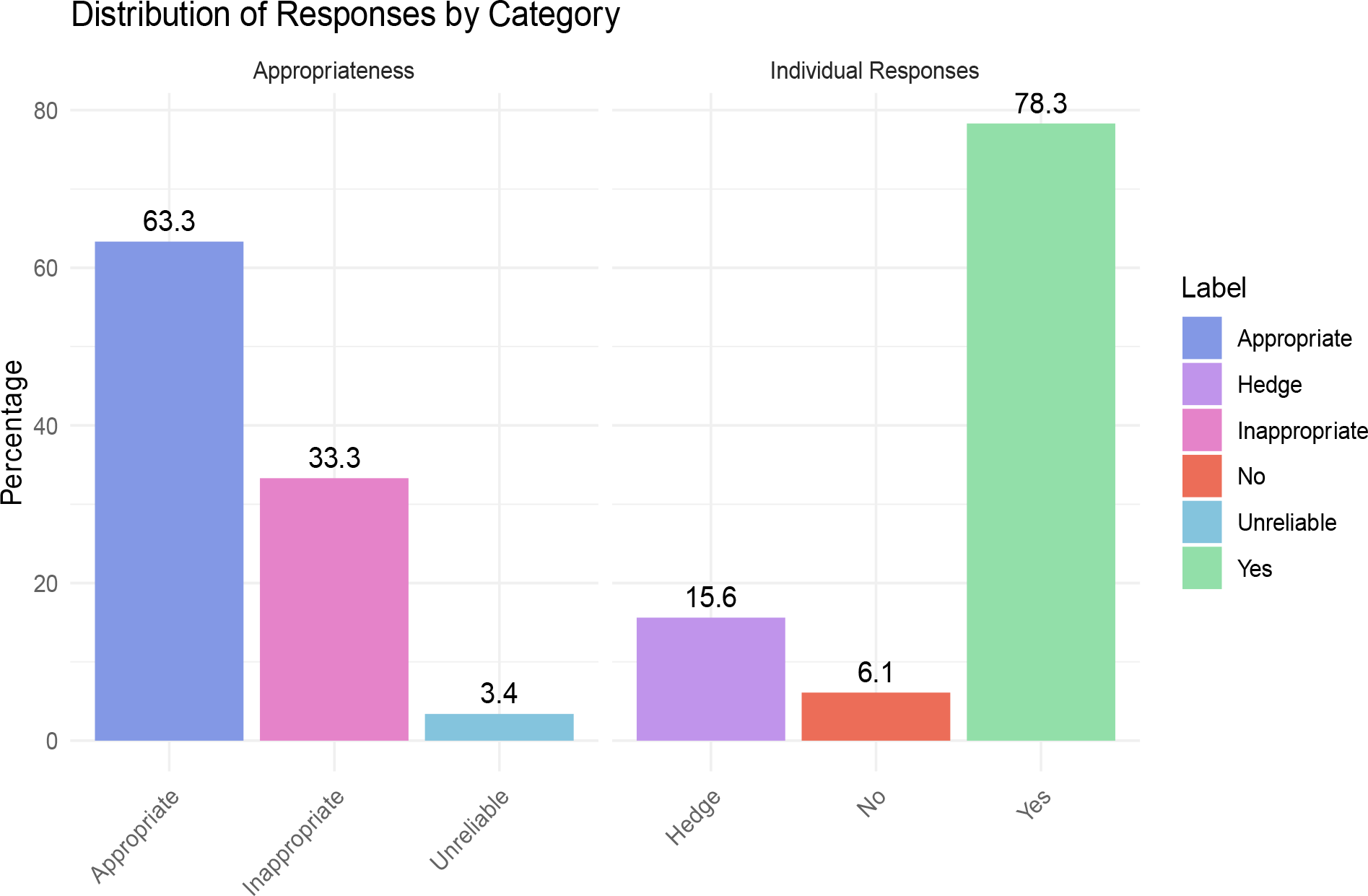
Percentage Assessment of Response Appropriateness Based on Graded Responses as Hedge, No and Yes. This figure presents the percentage-based assessment of response correctness and appropriateness. The horizontal axis represents the appropriateness scale and graded responses, while the vertical axis represents appropriateness percentages. Color coding is used to distinguish between different response categories, enhancing the clarity of the results.

**Figure 2.**
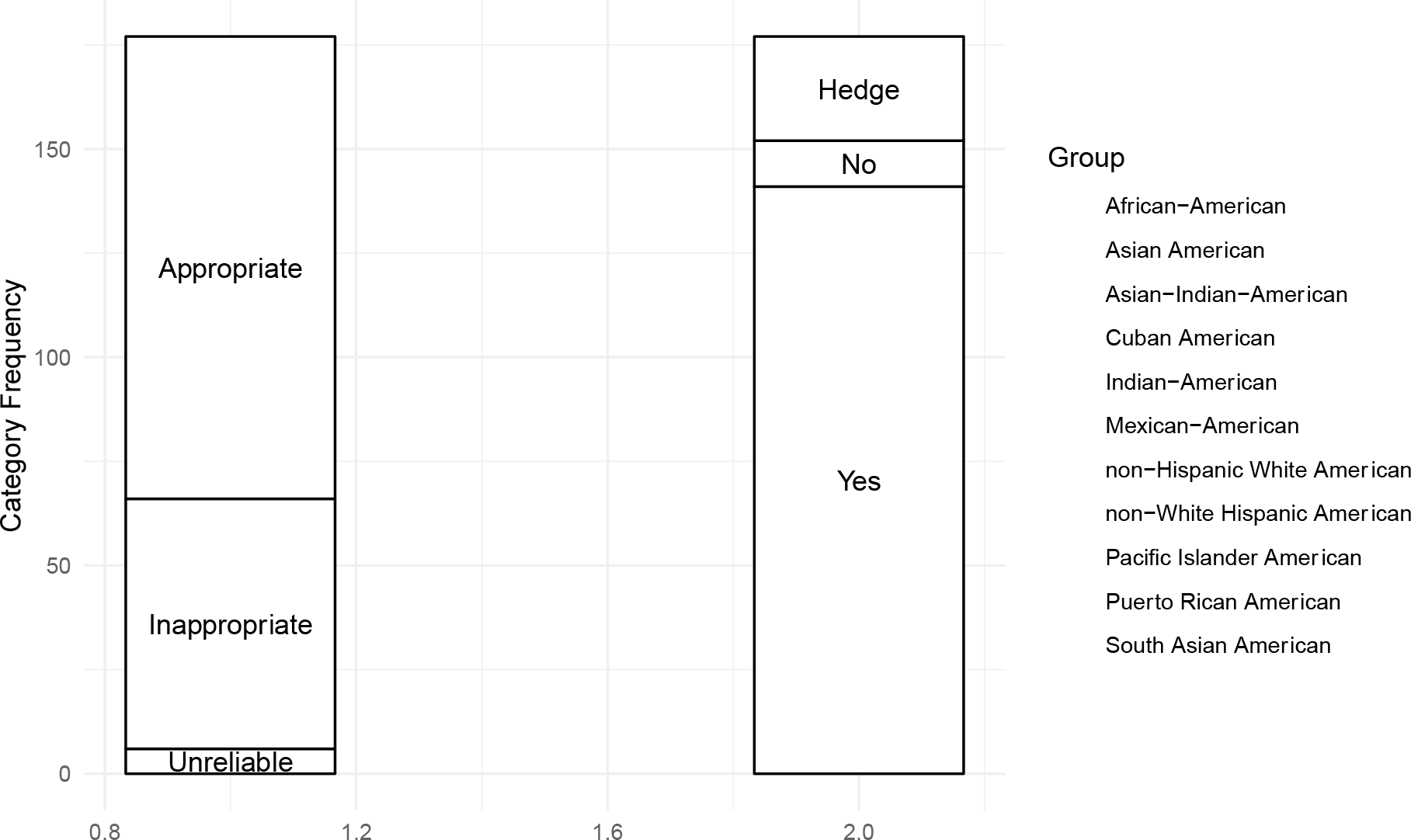
Alluvial Diagram Illustrating the Appropriateness of Responses with Racial Categories. This alluvial diagram visually represents the appropriateness of responses provided within racial categories, showcasing the flow and distribution of responses across different racial groups. Each stream within the diagram represents a specific racial category, with varying widths representing the number of responses. The thickness of the connecting ribbons between the racial categories signifies the degree of overlap or agreement in responses between these categories. The diagram provides insights into the patterns of response appropriateness and allows for a nuanced examination of how responses align or diverge across racial categories.

**Figure 3.**
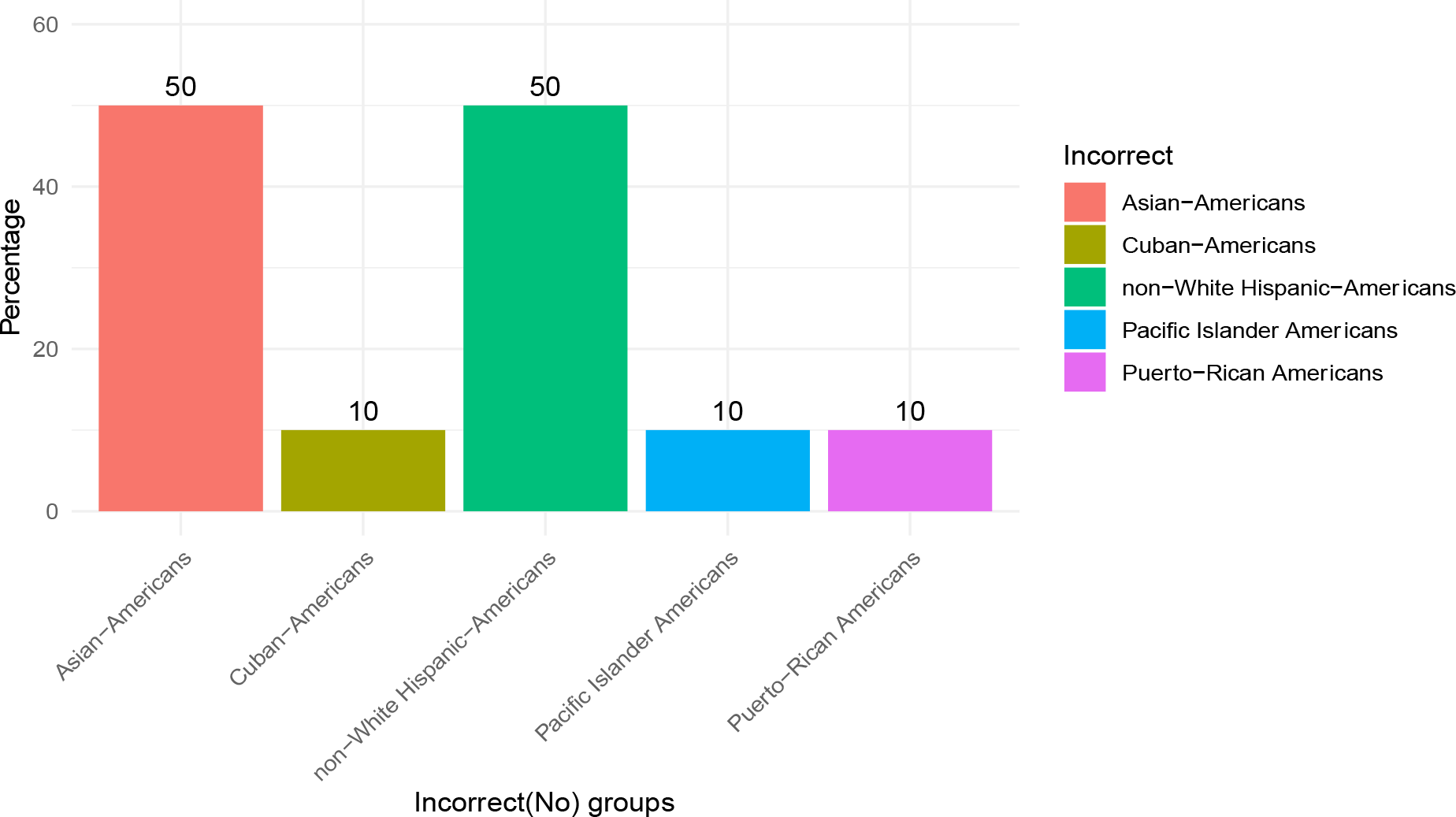
Bar Chart Illustrating Incorrect Responses by Racial Groups. This bar chart presents an analysis of incorrect responses grouped by racial categories. Each bar represents a specific racial group, and the height of each bar corresponds to the frequency or percentage of incorrect responses within that group. The chart provides a clear visual representation of how incorrect responses are distributed across different racial categories, offering insights into potential disparities or trends in response accuracy.

**Figure 4.**
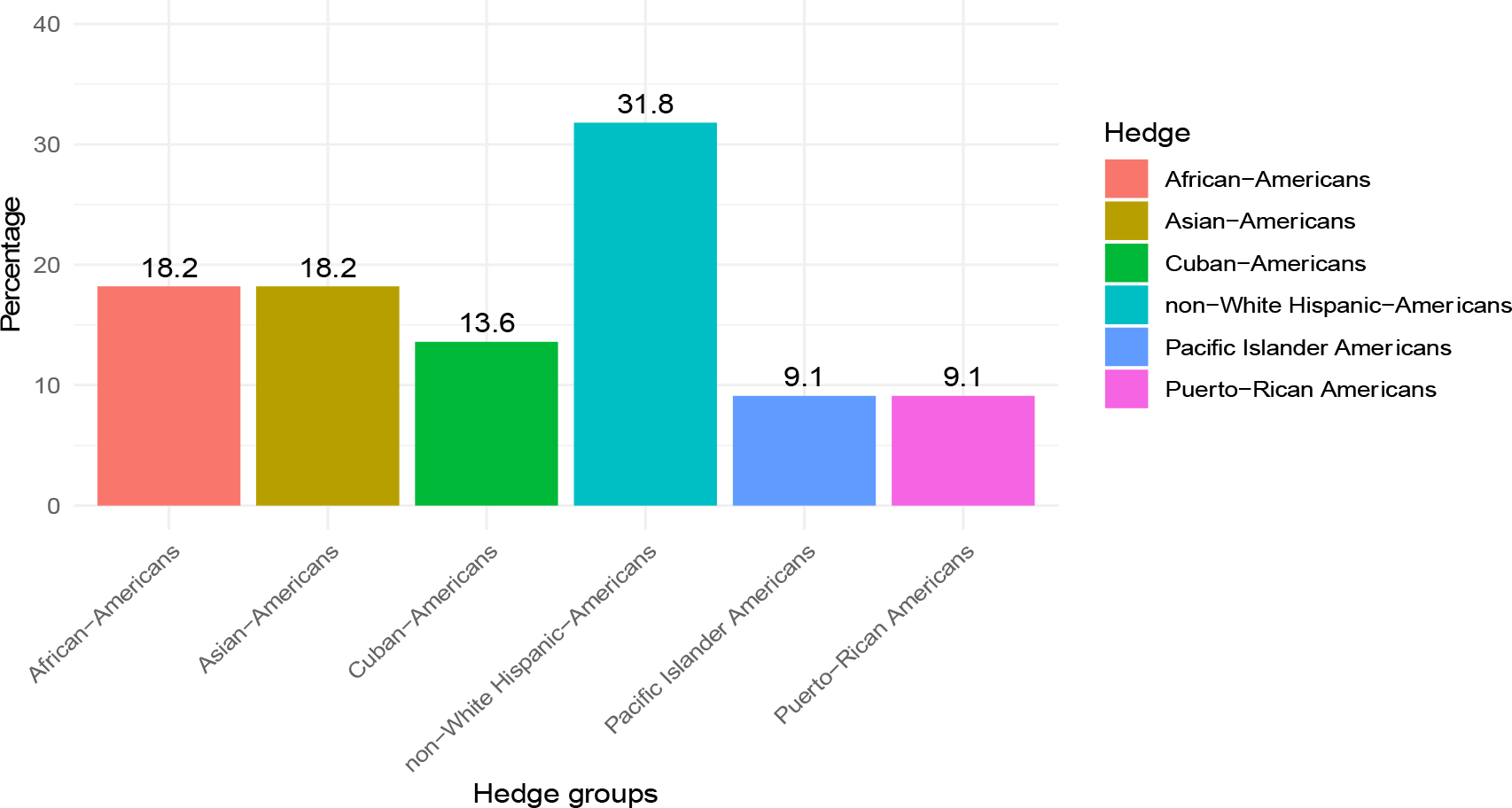
Bar Chart Displaying Indeterminate (Hedging) Responses by Racial Groups. This bar chart visually portrays the occurrence of indeterminate (hedging) responses among various racial groups. Each bar corresponds to a specific racial category, and the height of each bar represents the frequency or percentage of responses classified as indeterminate within that category.

**Figure 5.**
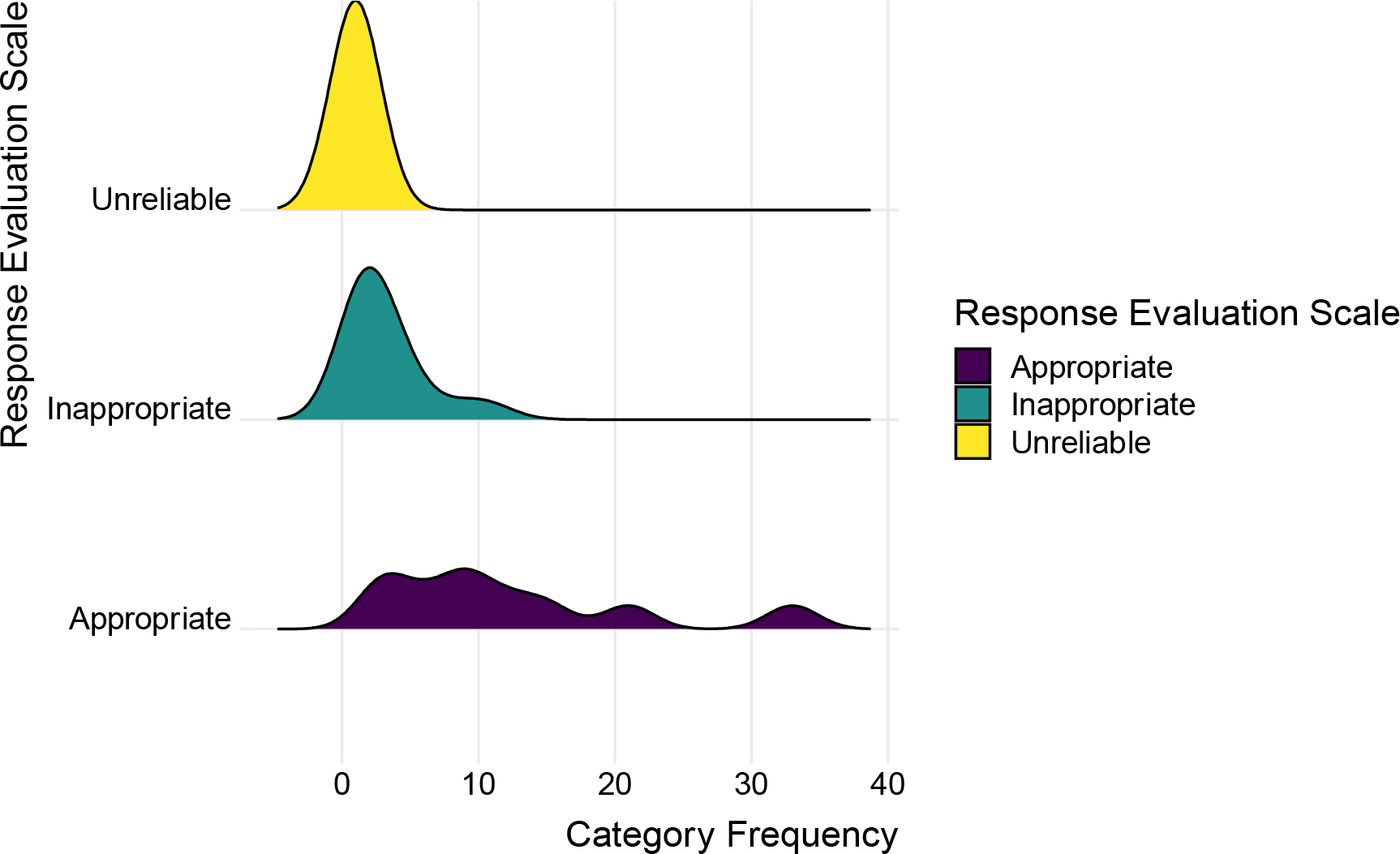
Distribution Curves of Response Appropriateness. This figure illustrates the distribution curves representing the assessment of response appropriateness using a three-color-coded evaluation scale. The three curves correspond to different categories: “Unreliable” (in yellow), “Inappropriate” (in green), and “Appropriate” (in purple), while in contrast, the “Appropriate” curve exhibits a normal distribution with a greater spread over the response category frequency, suggesting a predominant yet balanced distribution of responses rated as appropriate.

**Figure 6.**
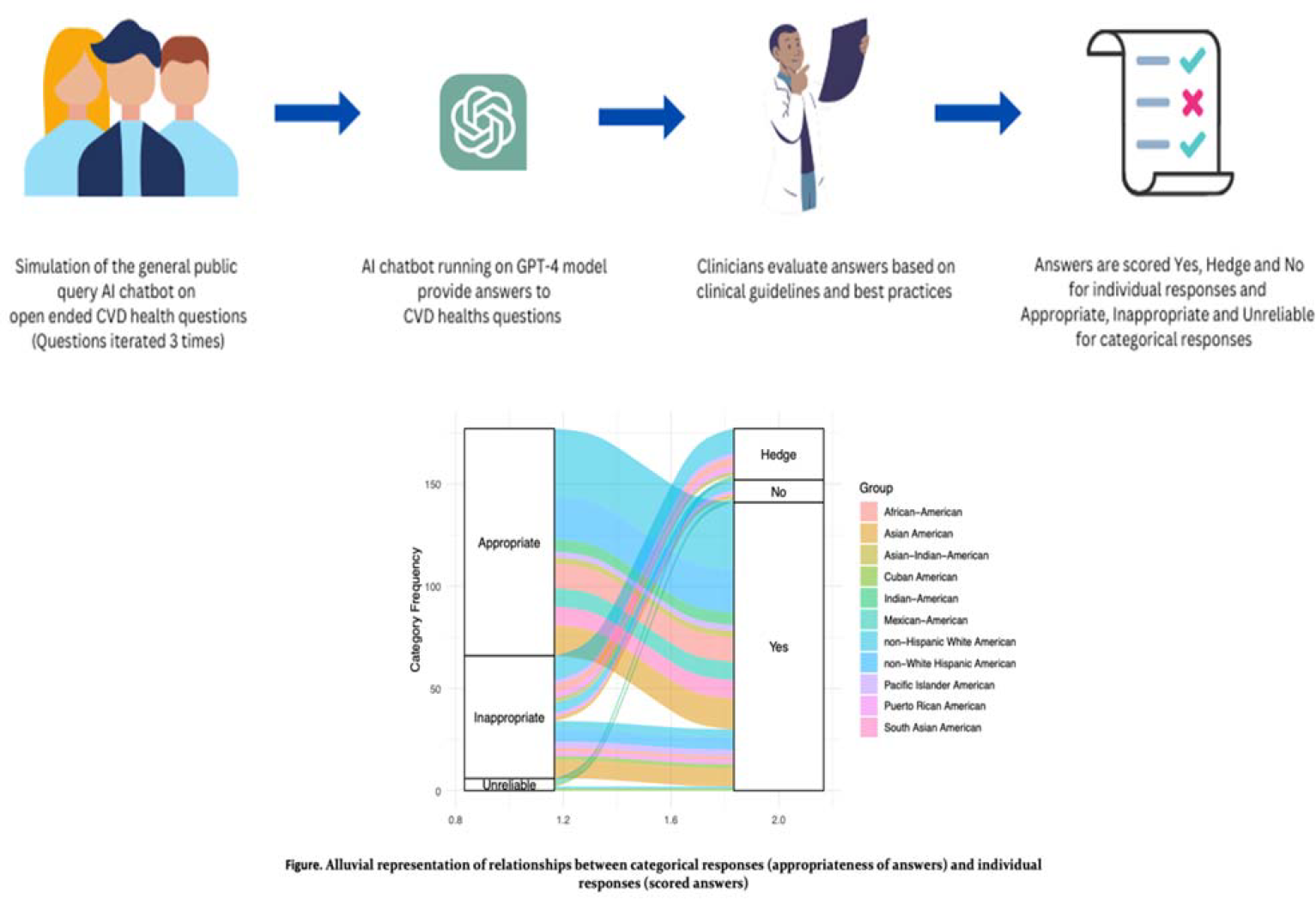

These findings further highlight the fact that AI-based online platform may sometimes further accentuate racial health disparities, underpinning the need for improved training of AI models on health disparities including cardiovascular racial disparities. ^22^

We found a consistent trend in which the most common responses, both incorrect and hedge, were related to non-White Hispanic Americans and Asian Americans. Another interesting finding is that while hedge responses from prompts related to African Americans who were the racial group with the second most common hedge responses at 18.2% (tie with Asian Americans), there were reassuringly no incorrect responses related to African Americans. Although the reason for this trend is unclear, potential explanations could be poor training of the AI-based platform on health disparities related to these groups, or a potential underrepresentation of these racial groups in health disparities research, such that the AI-platform has not available data to be trained on.

Findings from our study are important because with the widespread use of AI, effective programs for addressing health equity such working with community groups and directly addressing social determinants of health factors may be displaced by AI due to a perception of its superiority. ^12,23^ On account of this, every output from it may be regarded as correct, without necessarily being so, as shown by out study, hence worsening the health equity crisis. ^24^

Some limitations of our study include the fact that our study used a single AI model in assessing its knowledge of CVD racial disparities, as well as the fact that this online chat-based AI models are not optimized for medical use, as well as the fact that racial expression especially when applied in medicine is usually heterogenous.

### Summary

Our study showed that a popular online-based AI model has a satisfactory but suboptimal knowledge of CVD racial disparities. Given that these have far reaching consequences, including an accentuation of pre-existing CVD racial disparities, prompt action must be taken to better train AI model on health disparities.

## Data Availability

None

## Article Information

## Acknowledgements

None.

## SUPPLEMENTAL MATERIAL

### Supplemental Data

Direct output ChatGPT prompt responses are in separate supplemental documents as Answer Sets A, B, and C.

Topic Analysis and Associated Racial Disparity

#### Hedge

- *Heart disease* in non-Hispanic White Americans
- *Myocardial infarction* among non-Hispanic White and **Asian** Americans
- *Heart failure* among non-Hispanic White and **Pacific Islander** American **woman**
- High functional impairment from *acute coronary syndrome* among **Asian** and **non-White Hispanic** Americans
- Death rate from a*cute coronary syndrome* among **Asian** Americans
- Inherent *lipoprotein elevation* among non-Hispanic White and **African** Americans
- *Metabolic syndrome* development among **Cuban** Americans
- Inherent *triglyceride elevation* among **non-White Hispanic and African Americans**
- Likelihood of a higher *body mass index (BMI)* among **Cuban and Puerto Rican Americans**
- Development of *postpartum cardiomyopathy* among **non-White Hispanic Americans**

% of hedge responses involving disparities affecting a minority, or vulnerable group = 79% (22/28)

#### No

- *Stroke* in **Pacific Islanders**
- Death rate from *acute coronary syndrome* among **non-White Hispanic Americans**
- Death from *systemic hypertension* among **Puerto Rican Americans**
- *Blood pressure control* among **non-White Hispanic Americans**
- Likelihood of the development of *diabetes mellitus and peripheral vascular disease*
- among **Asian** Americans
- Risk of development of *metabolic syndrome* among **Cuban Americans**
- *Triglyceride levels* among n**on-White Hispanic American**

-% of no responses involving disparities affecting a minority, or vulnerable group = 91% (10/11).

### Key

**Racial sub-group with a hedge/no response**

*Cardiovascular disease/risk factor/related condition*

